# HLA-A*01:01 allele vanishing in COVID-19 patients population associated with non-structural epitope abundance in CD8^+^ T-cell repertoire

**DOI:** 10.1101/2022.07.05.22277214

**Authors:** Maxim Shkurnikov, Stepan Nersisyan, Darya Averinskaya, Milena Chekova, Fedor Polyakov, Aleksei Titov, Dmitriy Doroshenko, Valery Vechorko, Alexander Tonevitsky

## Abstract

In mid-2021, the SARS-CoV-2 Delta variant caused the third wave of the COVID-19 pandemic in several countries worldwide. The pivotal studies were aimed at studying changes in the efficiency of neutralizing antibodies to the spike protein. However, much less attention was paid to the T-cell response and the presentation of virus peptides by MHC-I molecules. In this study, we compared the features of the HLA-I genotype in symptomatic patients with COVID-19 in the first and third waves of the pandemic. As a result, we could identify the vanishing of carriers of the HLA-A*01:01 allele in the third wave and demonstrate the unique properties of this allele. Thus, HLA-A*01:01-binding immunoprevalent epitopes are mostly derived from ORF1ab. A set of epitopes from ORF1ab was tested, and their high immunogenicity was confirmed. Moreover, analysis of the results of single-cell phenotyping of T-cells in recovered patients showed that the predominant phenotype in HLA-A*01:01 carriers is central memory T-cells. The predominance of T-lymphocytes of this phenotype may contribute to forming long-term T-cell immunity in carriers of this allele. Our results can be the basis for highly effective vaccines based on ORF1ab peptides.

## Introduction

The Delta variant of SARS-CoV-2 caused the third wave of the COVID-19 pandemic in mid-2021 in many countries, including Russia (1). The surge in incidence was associated with the high transmissibility of this strain compared to the alpha variant (2). The increase in transmissibility mainly was due to the rise in the number of viral particles exhaled at the peak of infection by an infected person (up to 6 times compared to the Alpha variant) (3). Aside from the increased risk of hospitalization, the Delta variant also increases the risk of death in unvaccinated COVID-19 patients (4).

In the Delta variant, 18 protein level mutations significantly changed the course of the disease (5). Five were located in the Spike protein and significantly decreased the effectiveness of humoral immunity formed either naturally (recovered patients) or after vaccination (6). In addition, one of these mutations (D614G) increased the affinity of the receptor-binding domain (RBD) of the Spike protein to the ACE2 receptor (7). Finally, it should be noted that the rate of virus replication has also changed: the Delta variant replicates two times slower than the Alpha variant in the first 8 hours after infection *in vitro* (8). This circumstance is crucial since the non-structural protein ORF8 produced by SARS-CoV-2 can directly interact with major histocompatibility complex class I (MHC-I) molecules and suppress their maturation. As a result, the export of MHC-I molecules to an infected cell’s surface almost wholly stops after 18 hours of ORF8 expression (9).

MHC-I molecules are one of the key mediators of the first steps of a specific immune response to COVID-19. Right after entering a cell, SARS-CoV-2 induces the translation of its proteins. Some of these proteins enter the proteasomes of the infected cell, become cleaved to short peptides (8–12 amino acid residues), and bind to MHC-I molecules. After binding, the complex consisting of the MHC-I molecule and the peptide is transferred to the infected cell’s surface, where it can interact with the T-cell receptor (TCR) of CD8^+^ naïve, central memory or the effector memory T-cell subpopulations. In response to the interaction, the CD8^+^ T-lymphocyte destroys the infected cell using perforins and serine proteases (10). The crucial role of long-term CD8^+^ T-cell activation in the immune response to SARS-CoV-2 has been recently studied in a cohort of patients with different severity of disease (11,12).

There are three main types of MHC-I molecules encoded by HLA-A, HLA-B, and HLA-C (Human Leukocyte Antigens) genes. Each gene is presented in two variants (alleles) inherited from parents. There exist dozens of HLA alleles, and every allele encodes an MHC-I molecule with an individual ability to bind various self- and non-self-antigens (13). Individual combinations of HLA class I alleles essentially associated with the severity of multiple infectious diseases, including malaria (14), tuberculosis (15), HIV (16), and viral hepatitis (13). Previously we have demonstrated the important role of MHC-I peptide presentation in the development of a specific immune response to the Wuhan-Hu-1 variant (GISAID accession EPI_ISL_402125) (17).

For over two years of the COVID-19 pandemic, the scientific community accumulated a significant amount of information on the actual epitopes of various SARS-CoV-2 variants (18), features of the formation of T-cell memory (19), trends in the frequency of mutations in the virus (20). However, the associations of the HLA genotypes and the course of COVID-19 were mainly analyzed according to data related to the first wave of the pandemic and the initial virus variant (21–24). Moreover, in studies of the associations between the severity of COVID-19 and HLA-I genotypes, the age of the recovered patients was practically not considered. It should be noted that age is a significant factor in the immune response to COVID-19 (25–27). In particular, it has been shown that in people over 60 years of age, telomere lengths of naïve T-lymphocytes decrease significantly, which leads to an almost 10-fold drop in their ability to divide upon activation (28). Also, the T-cell receptor (TCR) repertoire is reduced in older people (29).

Previously, in a cohort of the first wave of COVID-19 patients, we showed that the low number of peptides presented by an individual’s HLA-I genotype significantly correlates with COVID-19 severity only in patients under the age of 60 (17). In this study, we compared the features of the HLA-I genotypes of COVID-19 patients under 60 between the first and the third waves. In addition, we assessed the influence of mutations in the SARS-CoV-2 variants on the immunogenic epitopes of CD8^+^ T-lymphocytes.

## Materials and methods

### Design and Participants

Three groups of patients were enrolled in the study. First, the control group of 428 volunteers was established using electronic HLA genotype records of the Federal Register of Bone Marrow Donors (Pirogov Russian National Research Medical University) (17). The group of 147 patients of the first wave of COVID-19 was formed from May to August 2020 (Wave 1 group). Out of them, the subset of 28 COVID-19 convalescent donors of the first wave was enrolled in a prospective trial (CPS group). Finally, the group of patients of the third wave was formed from June to July 2021 (Wave 3 group). 219 COVID-19 patients in Wave 3 group enrolled in O.M. Filatov City Clinical Hospital, (Moscow, Russia).

All patients had at least one positive test result for SARS-CoV-2 by reverse transcription PCR (RT-qPCR) from nasopharyngeal swabs or bronchoalveolar lavage. Patients with pathologies that led to greater morbidity or who had additional immunosuppression (patients with diabetes, HIV, active cancer in treatment with chemotherapy, immunodeficiency, autoimmune diseases with immunosuppressants, and transplants) were not included in the study. The medical practitioner collected blood (2 ml) in ethylenediaminetetraacetic acid (EDTA) tube for HLA genotyping.

The severity of the disease was defined according to the classification scheme used by the US National Institutes of Health (from www.covid19treatmentguidelines.nih.gov): asymptomatic (lack of symptoms), mild severity (fever, cough, muscle pain, but without respiratory difficulty or abnormal chest imaging) and moderate/severe (lower respiratory disease at CT scan or clinical assessment, oxygen saturation (SaO2) > 93% on room air, but lung infiltrates less than 50%).

The study protocol was reviewed and approved by the Local Ethics Committee at the Pirogov Russian National Research Medical University (Meeting No. 194 of March 16, 2020, Protocol No. 2020/07), by the Local Ethics Committee at O.M. Filatov City Clinical Hospital (Protocol No. 237 of June 25, 2021), and by the National Research Center for Hematology ethical committee (N 150, 02.07.2020); the study was conducted in accordance with the Declaration of Helsinki.

### Human Leukocyte Antigen Class I Genotyping With Targeted Next-Generation Sequencing

Genomic DNA was isolated from the frozen collected anticoagulated whole blood samples using the QIAamp DNA Blood Mini Kit (QIAGEN GmbH, Hilden, Germany). HLA genotyping was performed with the HLA-Expert kit (DNA Technology LLC, Russia) by amplifying exons 2 and 3 of the HLA-A/B/C genes and exon 2 of the HLA-DRB1/3/4/5/DQB1/DPB1 genes. Prepared libraries were run on Illumina MiSeq sequencer using a standard flow-cell with 2х250 paired-end sequencing. Reads were analyzed using HLA-Expert Software (DNA-Technology LLC, Russia) and the IPD-IMGT/HLA database 3.41.0 (30).

HLA-genotyping of convalescent donors was performed using the One Lambda ALLType kit (Thermo Fisher Scientific, USA), which uses multiplex PCR to amplify full HLA-A/B/C gene sequences, and from exon 2 to the 30 UTR of the HLA525 DRB1/3/4/5/DQB1 genes as described previously (11). Prepared libraries were run on Illumina MiSeq sequencer using a standard flow-cell with 2х150 paired-end sequencing. Reads were analyzed using One Lambda HLA TypeStream Visual Software (TSV), version 2.0.0.27232, and the IPD IMGT/HLA database 3.39.0.0 (30). Processed genotype data are available in Supplementary Table S1.

### Peripheral blood mononuclear cell (PBMC) isolation

30 mL of venous blood from CPS group donors was collected into EDTA tubes (Sarstedt) and subjected to Ficoll (Paneco) density gradient centrifugation (400 x g, 30 min). Isolated PBMCs were washed with PBS containing 2 mM EDTA and used for assays or frozen in fetal bovine serum containing 7% DMSO.

### T-cell expansion

Full details of the T-cell expansion are provided in the manuscript (11). Briefly, PBMCs sampled form COVID-19 convalescents were expanded for 12 days with the pre-selected 94 SARS-CoV-2 peptides (final concentration of each = 10 μM). On days 10 and 11, an aliquot of cell suspension was used for anti–IFN-γ ELISA aiming at the identification of responses to individual peptides.

### Cell stimulation with individual peptides

After 10 days of expansion, an aliquot of cell culture was washed twice in 1.5 mL of PBS and was then transferred to AIM-V medium (Thermo Fisher Scientific), plated at 1 × 10e5 cells per well in 96-well plates, and incubated overnight (12–16 hours) with the smaller pools of peptides, spanning the composition of the initial peptide set. The following day the culture medium was collected and tested for IFN-γ as described below. If the cells reacted positively to one or several pools we sampled an aliquot of cell culture again on days 11–12, and stimulated it as described above individually with each peptide (2 μM) from those peptide pools. Only peptides predicted to bind to each individual’s HLA were tested.

### Anti–IFN-γ ELISA

Culture 96-well plates with cells, incubated with peptide pools or individual peptides were centrifuged for 3 minutes at 700g, and 100 μL of the medium was transferred to the ELISA plates and the detection of IFN-γ was performed (11). OD was measured at 450 nm on a MultiScan FC (Thermo Fisher Scientific) instrument (OD450).

Test wells (medium from cells, incubated with peptides) were compared with negative control wells (cells incubated with the solvent for peptides). Test wells with the ratio OD450_test_well/OD450_negative control ≥ 1.25 and the difference OD450_test_well – OD450_negative control ≥ 0.08 were considered positive. Peptides with a ratio between 1.25 and 1.5 were tested again up to 3 times as biological replicates to ensure the accuracy of their response, and peptides with 2 or 3 positive results were considered positive.

### Peptides

Peptides (at least 95% purity) were synthesized either by Peptide 2.0 Inc. or by the Shemyakin-Ovchinnikov Institute of Bioorganic Chemistry RAS.

### Assessing immunoprevalent epitopes for HLA-A*01:01 and HLA-A*02:01 and mutations in VOC

To assess the immunoprevalent epitopes of HLA-A*01:01 and HLA-A*02:01, we queried IEDB (18) for epitopes with positive MHC binding and positive T-cell assays using “Severe acute respiratory syndrome-related coronavirus” as “Organism” on May 17, 2022. Epitopes with more than 50% response rate for the respective allele carriers were considered immunoprevalent. Mutation analysis was performed using data extracted from T-cell COVID-19 Atlas (20) (accessed May 30, 2022) and included 16 VOC strains.

### Phenotype analysis of CD8+ T-lymphocytes of convalescent HLA-A*01:01 and HLA-A*02:01 alleles carriers

The data were obtained from supplementary materials of Francis et al. (19). The experiment conducted by the authors consisted in performing single-cell RNA sequencing (scRNA-seq) of T-cells bound to DNA-barcoded peptide-HLA tetramers. This approach allowed us to extract information on both TCR sequences and their specific HLA-epitope pairs. Francis with coauthors used a comprehensive library of SARS-CoV-2 epitopes and epitopes emerging from SARS-CoV-1, cytomegalovirus, Epstein–Barr virus, and influenza. For the analysis of the distribution of epitopes across the SARS-CoV-2 genome, the data file S3 was used. It includes the list of SARS-CoV-2 reactive clonotypes with their specific epitopes and their antigen source. In addition, the list of SARS-CoV-2 epitopes was extracted, and duplicate entries were removed.

The data file S7 was used for the analysis of the T-cell phenotypes. It comprises the information about the phenotypes of T-cells reacting to particular epitopes. The phenotypes of T-cells were computationally assigned by the authors based on the analysis of differentially expressed genes. The authors note that they identified several distinct cell clusters, but except for naïve cells, central memory, and fully activated cytotoxic effectors, all other clusters had mixed properties, making it impossible to determine their phenotypes univocally. Therefore, we decided to take only the definitely determined cell phenotypes into our analysis: naïve cells, central memory, and fully activated cytotoxic effector cells.

### Bioinformatics analysis of HLA/peptide interactions

Protein sequences of SARS-CoV-2 variants were obtained from GISAID portal (Elbe & Buckland-Merrett, 2017): EPI_ISL_402125 (Wuhan), EPI_ISL_1663516 (Delta, B.1.617.2), EPI_ISL_6699752 (Omicron BA.1, B.1.1.529), EPI_ISL_9884589 (Omicron BA.2, B.1.1.529), EPI_ISL_9854919 (Omicron BA.3, B.1.1.529), EPI_ISL_11873073 (Omicron BA.4, B.1.1.529). T-CoV pipeline was executed to analyze HLA/peptide interactions (5,20). In the pipeline’s core, binding affinities of all viral peptides and 169 frequent HLA class I molecules were predicted using NetMHCpan 4.1 (31).

### Statistical Analysis

Allele frequencies in considered cohorts were estimated by dividing the number of occurrences of a given allele in individuals by the doubled total number of individuals (i.e., identical alleles of homozygous individuals were counted as two occurrences). The following functions from the stats library in R were used to conduct statistical testing: fisher.test for Fisher’s exact test, wilcox.test for Mann-Whitney U test. In addition, the Benjamini-Hochberg procedure was used to perform multiple testing corrections. Plots were constructed with ggpubr and pheatmap libraries.

## Results

### Distribution of HLA Class I Alleles in the cohorts of patients of the first and the third waves of COVID-19

We performed HLA class I genotyping of 147 patients who tested positive for COVID-19 during the first wave of COVID-19 in Moscow (from May to August 2020, Wave 1 group). Also, 219 COVID-19 patients were genotyped from June to July 2021 (Wave 3 group). The demographic and clinical data of these groups are summarized in Table 1. We did not find significant differences in the age of patients in the comparison groups and the gender ratio in the groups. The fraction of vaccinated patients (two doses of Sputnik V vaccine) in the Wave 3 group was insignificant and equal to 8.7%, slightly lower than the citywide vaccination rate for June 2021 – 15% (32). There was a significant increase in the proportion of patients with obstructive pulmonary disease (Fisher’s exact test p = 0.01), obesity (Fisher’s exact test p = 0.01, OR = 3.8), hypertension (Fisher’s exact test p = 0.003, OR = 2.4) in the third wave of COVID-19. We also assessed the contribution of comorbidities to the risk of death from COVID-19. Interestingly, heart disease hypertension (Fisher’s exact test p = 5.2е-5, OR = 13) and hypertension (Fisher’s exact test p = 0.045, OR = 3.5) were significant death risk factors in the first wave. At the same time, no similar effects were observed for the third wave patients (Supplementary Table S2).

**Table 1.** Characteristics of participants by groups.

|  | Wave 1 | Wave 3 | p-value |
| --- | --- | --- | --- |
| **n** | 147 | 219 |  |
| **Age** | 43.4 +/- 11 | 42.9 +/- 5.2 | 0.4 |
| **Sex** |  |  |  |
| male | 73 (49.7%) | 123 (56.2%) | 0.24, OR = 0.77 |
| female | 74 (50.3%) | 96 (43.8%) |  |
| **Vaccination** | 0 | 19 (8.7%) | 4.9e-05 |
| **Severity** |  |  |  |
| mild/moderate | 120 (81.6%) | 192 (87.7%) | 0.13, OR = 0.63 |
| severe | 27 (18.4%) | 27 (12.3%) |  |
| **Outcome** |  |  |  |
| recovered | 127 (86.4%) | 197 (90%) | 0.32, OR = 0.71 |
| deceased | 20 (13.6%) | 22 (10%) |  |
| **ICU** | 28 (19%) | 51 (23.3%) | 0.36, OR = 1.3 |
| **Oxygen support** |  |  |  |
| non | 98 (66.7%) | 140 (63.9%) | 8.8e-05 |
| low flow nasal oxygen supplementation | 24 (16.3%) | 38 (17.4%) |  |
| prone position | 0 | 11 (5%) |  |
| high-flow oxygen therapy | 16 (10.9%) | 4 (1.8%) |  |
| mechanical ventilation | 9 (6.1%) | 26 (11.9%) |  |
| **Obstructive pulmonary disease** | 0 | 9 (4.1%) | 0.01, OR = inf |
| **Obesity** | 4 (2.7%) | 21 (9.6%) | 0.01, OR = 3.8 |
| **Heart diseases** | 20 (9.1%) | 14 (9.5%) | 1, OR = 0.95 |
| **Hypertension** | 16 (10.9%) | 50 (22.8%) | 0.003, OR = 2.4 |
| **Neoplasma** | 3 (2%) | 9 (4.1%) | 0.4, OR = 2.1 |

First, we tested whether the frequency of a single allele can differentiate individuals from three groups: COVID-19 patients of the first wave, COVID-19 patients of the third wave, and the control group. The distribution of major HLA-A, HLA-B, and HLA-C alleles in these three groups is summarized in Figure 1. Fisher’s exact test was used to make formal statistical comparisons. As a result, we found that for all possible group comparisons, only one allele out of dozen top alleles had a high odds ratio, which can be considered statistically significant after multiple testing correction. Specifically, frequency of HLA-A*01:01 allele decreased from 17.3% in the Wave 1 group to 9.8% in the Wave 3 group (Fisher’s exact test adj.p = 0.025, OR = 0.5). Some of the alleles were differentially enriched if no multiple testing correction was applied (Supplementary Table S3).

**Figure 1.**
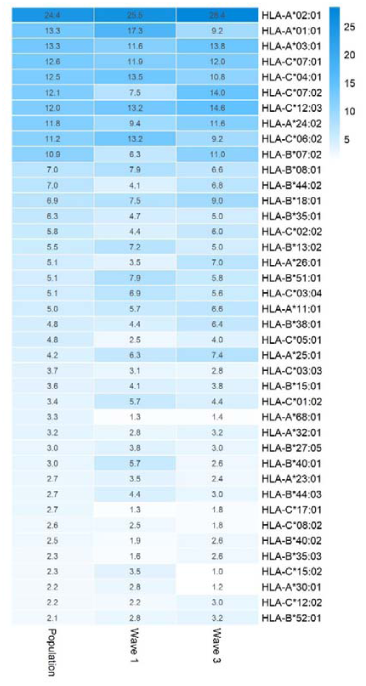
Distribution of HLA-A, HLA-B, and HLA-C allele frequencies in the groups of COVID-19 patients and the control group. Alleles with frequency over 5% in at least one of three considered groups are presented.

We hypothesized that the decrease in the frequency of HLA-A*01:01 carriers among patients of the third wave group could be attributed to the characteristics of the T-cell response. To test this hypothesis, computational methods were used to analyze the interactions between the HLA-I molecules and the viral peptides, the effects of mutations in variants of concern (VOC) on these interactions, as well as the results of experimental testing of the immunogenicity of peptides in patients who recovered from the COVID-19 in the first wave.

### Analysis of the binding affinity of SARS-CoV-2 peptides with MHC-I molecules

The nonstructural proteins encoded by ORF1ab gene are translated first (33) and undergo proteasomal digestion before structural and accessory proteins (including ORF8 which suppress MHC-I maturation). Given that, we analyzed the ability of MHC-I molecules encoded by 12 most common alleles in the European population (34) (Figure 2) to bind and present ORF1ab and non-ORF1ab peptides. The ability of HLA-A*01:01 to interact with peptides of both structural and non-structural proteins of the virus was moderate. Namely, it was predicted to interact with an affinity of less than 50 nM (tight binding peptides) with 28 peptides from ORF1ab and 8 peptides from structural and accessory proteins. Another allele, HLA-A*02:01, was the most frequent in the population and had 207 predicted tight binders from ORF1ab and 56 tight binders from the other proteins. We also compared the ability to present viral peptides between genotypes that include HLA-A*01:01 and genotypes which do not include this allele (Figure S1 A). HLA-I genotypes, which included HLA-A*01:01, had, on average, a lower number of high-affinity peptides compared to non-carriers regardless of wave (Mann-Whitney U test p < 0.02).

**Figure 2.**
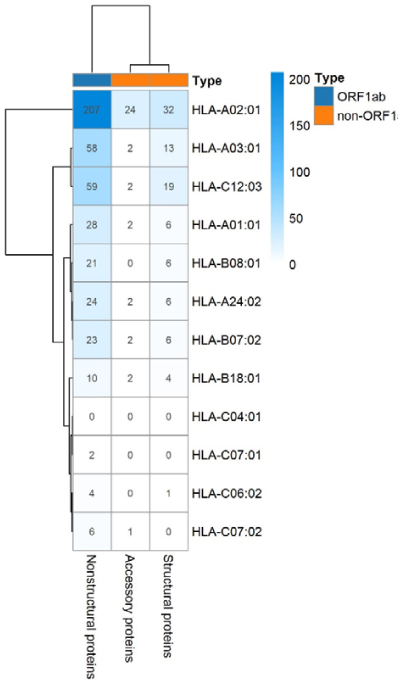
The number of virus peptides interacting with the most frequent alleles with an affinity of no more than 50 nM.

It should be noted that variant AY.122 (1) prevailed in the third wave of COVID-19 in the Moscow region. One of its characteristic mutations is G8R in the NS8 (ORF8) protein. This mutation causes a decrease in the affinity of the interaction of peptides NS8 _5-13_ (FLGIITTV) and NS8 _2-13_ (FLVFLGIITTV) with the MHC-I molecule encoded by the most frequent HLA-A*02:01 allele for the analyzed population. This mutation is associated with a non-significant reduction in the number of high-affinity accessory protein epitopes in HLA-A01:01 carriers (Figure S1 B).

### Analysis of immunogenic epitopes

Next, we analyzed the immunogenic epitopes of SARS-CoV-2 using the data from the IEDB portal for HLA-A*01:01 and HLA-A*02:01 alleles. At the time of the analysis (May 2022), the database contained information on the T-cell responses to 365 viral peptides. Two shared epitopes were found for both alleles: S _136-144_ (CNDPFLGVY), M _170-178_ (VATSRTLSY). For the HLA-A*01:01 allele, the immunogenicity of 50 ORF1ab peptides and 33 non-ORF1ab peptides was validated. In entire agreement with computational predictions, the number of epitopes was higher for the HLA-A*02:01 allele: 139 ORF1ab peptides and 145 non-ORF1ab peptides. The ratio of peptides from ORF1ab for the HLA-A*01:01 allele was 1.6 times higher than for the HLA-A*02:01 allele (Fisher’s Exact Test p-value = 0.08). Among the tested peptides, immunoprevalent epitopes were identified (i.e., caused a response of T-lymphocytes in at least 50% of the tested samples). For the HLA-A*01:01 allele, 10 immunoprevalent epitopes from ORF1ab and only 3 epitopes not from ORF1ab were identified. For the HLA-A*02:01 allele, 51 immunoprevalent epitopes from ORF1ab and 59 from non-ORF1ab were identified. The ratio of immunoprevalent epitopes from ORF1ab for the HLA-A*01:01 allele was 3.8 times higher than for the HLA-A*02:01 allele (Fisher’s Exact Test p-value = 0.04).

### Validation of ORF1ab epitopes in a subset of convalescent HLA-A*01:01 and HLA-A*02:01 carriers

To validate the hypothesis that the first wave convalescent HLA-A*01:01 allele carriers had a high number of immunogenic epitopes from ORF1ab proteins, we analyzed the T-cell responses of 28 patients with the history of confirmed COVID-19 during first wave who carried at least one of the two most common alleles in the European population: HLA-A*01:01 (13 patients) and HLA-A02:01 (15 patients). Individual immunogenicity of 15 validated epitopes from SARS-CoV-2 ORF1ab (Supplementary Table S4) was tested for each T-cell sample. The peptides from this panel did not induce a T-cell response in patients who had not previously had COVID-19 (11). The number of epitopes for HLA-A*01:01 was 7 for HLA-A*02:01 – 8.

Since time from the symptoms onset to the first measurement could affect the strength of the T-cell response, we compared these times between groups of HLA-A*01:01 and HLA-A*02:01 carriers. For HLA-A*01:01 carriers, the median time after symptoms onset was 34 days, and for HLA-A*02:01 carriers, it was 41 days; the differences were insignificant (Figure 3A). At the same time, the true positive rates for these alleles differed significantly for epitopes from ORF1ab (Figure 3B). Three of the 7 HLA-A*01:01 epitopes tested were immunoprevalent (ORF1ab _1637-1646_ TTDPSFLGRY, ORF1ab _1636-1646_ HTTDPSFLGRY, ORF1ab _1321-1329_ PTDNYITTY), but there were no immunoprevalent epitopes out of 8 tested for HLA-A*02:01.

**Figure 3.**
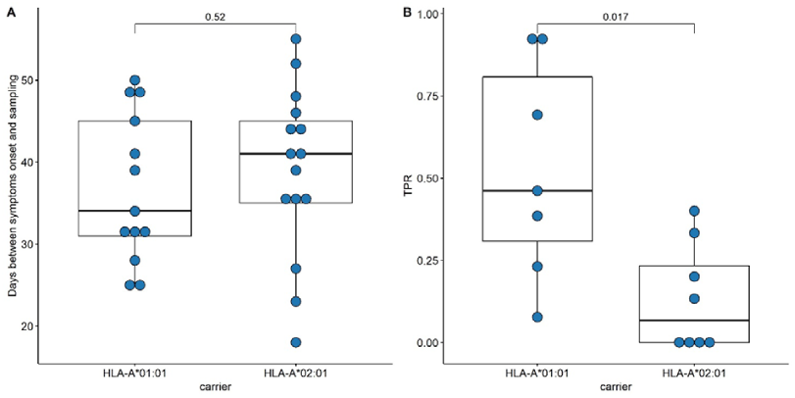
Validation of ORF1ab epitopes immunogenicity in a subset of convalescent HLA-A*01:01 and HLA-A*02:01 carriers. (A) Distribution of days between symptoms onset and blood sampling in the comparison groups. (B) True positive rates for the ORF1ab epitope set in the comparison groups.

### Phenotype analysis of CD8+ T-lymphocytes of convalescent patients

Aside from IEDB and own epitope validation data, we analyzed *ex vivo* scRNA-seq data of convalescent individuals CD8^+^ T-cells activated by single peptides (19). The considered dataset included the set of phenotyped T-cell clones responding to a comprehensive set of SARS-CoV-2 derived epitopes associated with four major HLA class I alleles (HLA-A*01:01, HLA-A*02:01, HLA-A*24:02, and HLA-B*07:02). First, we examined the distribution of SARS-CoV-2 derived epitopes which elicited T-cell response according to the genomic region from which they originated. Concordantly with the results mentioned above, the majority of HLA-A*01:01 epitopes were from ORF1ab. The same tendency was not observed for the other alleles (Figure 4A).

**Figure 4.**
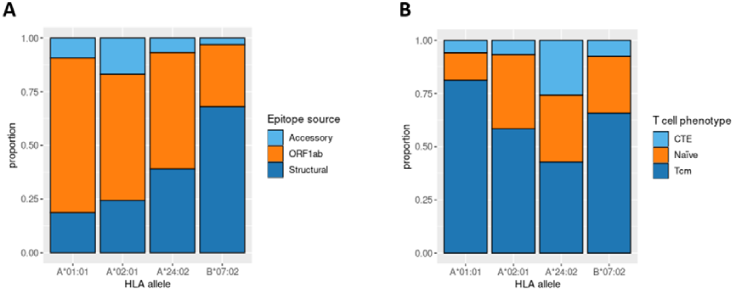
Phenotype analysis of CD8+ T-lymphocytes of convalescent patients. (A) The distribution of SARS-CoV-2 derived immunogenic epitopes according to their genomic origin. Epitopes associated with the HLA-A*01:01 allele tend to originate mainly from the conservative ORF1ab region, suggesting a prevalence of memory T-cells among the responding T-cells during the second encounter with SARS-CoV-2. (B) The ratio of responding T-cell phenotypes to the comprehensive set of SARS-CoV-2 derived epitopes. Tcm stands for T central memory subpopulation, and CTE stands for cytotoxic T effector cells. Epitopes associated with the HLA-A*01:01 allele elicit a more robust Tcm response compared with other alleles (pairwise Fisher exact test values: 5.423e-05 for A*01:01 against A*02:02 comparison, 0.0178 for A*01:01 against B*07:02 comparison, 6.196e-05 for A*01:01 against A*24:02 comparison).

Next, we analyzed the phenotypes of responding T-cell clones (Figure 4B). HLA-A*01:01-associated epitopes elicited responses mainly from the Tcm subpopulation. At the same time, the proportion of Tcm responding cells for other alleles was significantly smaller (Fisher Exact Test pairwise comparisons of A*01:01 with A*02:01, B*07:02, and A*24:02 results in p-values < 0.02). Together with the observation that most of the known HLA-A*01:01-associated epitopes originated from the conservative ORF1ab region, these results imply that people bearing HLA-A*01:01 may have a higher chance of eliciting robust immune response upon secondary exposure to SARS-CoV-2 due to the pre-existing immune memory.

### Analysis of the mutations in the VOC on CD8^+^ epitopes

Then, we analyzed the changes in the affinity of the interactions of immunoprevalent epitopes with HLA-A*01:01 and HLA-A*02:01 induced by mutations in VOCs, including the Delta variant. Strikingly, none of the HLA-A*01:01 immunoprevalent epitopes were affected by mutations in the current VOCs: Delta G/478K.V1 and Omicron (BA.1 – BA.4). At the same time, seven HLA-A*02:01 immunoprevalent epitopes were affected by the mutations. Six were located in the Spike protein and one in the NS3 protein. All except one mutation led to the decrease in the affinity of the interaction with HLA-A*02:01. Specifically, in Omicron BA.1 the S _495-503_ YGFQPTNGV peptide had three substitutions (G496S, Q498R, N501Y), which resulted in an increase in the affinity of its interaction with HLA-A*02:01 (from 2160 nM to 87 nM). In VOC Omicron BA.1 and BA.3, A67V and HV 69-70 deletion mutations resulted in the disappearance of the immunoprevalent epitope S _62-70_ VTWFHAIHV.

In addition, the effect of mutations in the VOCs on the binding affinity of all possible viral peptides with the 12 abovementioned HLA-I alleles was analyzed (Table 2). HLA-A*01:01 had 51 high-affinity peptides from ORF1ab and 13 high-affinity peptides from other proteins (structural and accessory) for the Wuhan-Hu-1 strain. A distinctive feature of this allele was that it had a relatively high number of high-affinity peptides, although VOC mutations do not affect them (Figure 5). Of the 64 high-affinity HLA-A*01:01 peptides, only one peptide significantly changed the affinity in 16 VOCs analyzed. At the same time, similar to the number of tight binders allele HLA-A*24:02 had 27 altered peptides from ORF1ab (Fisher’s exact test p-value = 3e-09, OR = 0.02). It should be noted that the high-affinity peptides from ORF1ab for all analyzed alleles were less affected by the mutations compared to the rest of the peptides (Mann-Whitney U test p-value = 4.3e-05).

**Table 2.**
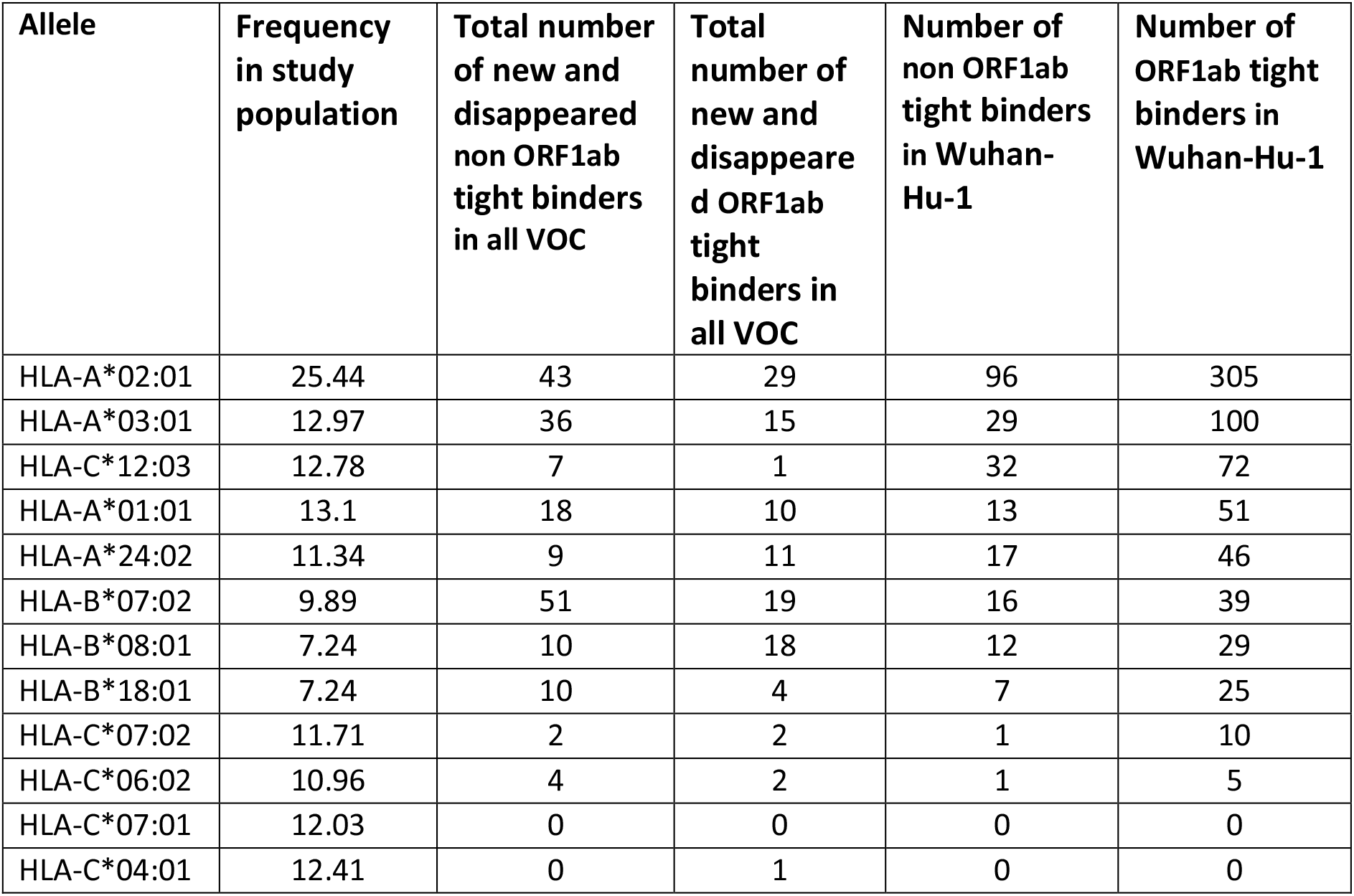
The number of high-affinity peptides mutated in VOCs.

| Allele | Frequency in study population | Total number of new and disappeared non ORF1ab tight binders in all VOC | Total number of new and disappeared ORF1ab tight binders in all VOC | Number of non ORF1ab tight binders in Wuhan-Hu-1 | Number of ORF1ab tight binders in Wuhan-Hu-1 |
| --- | --- | --- | --- | --- | --- |
| HLA-A*02:01 | 25.44 | 43 | 29 | 96 | 305 |
| HLA-A*03:01 | 12.97 | 36 | 15 | 29 | 100 |
| HLA-C*12:03 | 12.78 | 7 | 1 | 32 | 72 |
| HLA-A*01:01 | 13.1 | 18 | 10 | 13 | 51 |
| HLA-A*24:02 | 11.34 | 9 | 11 | 17 | 46 |
| HLA-B*07:02 | 9.89 | 51 | 19 | 16 | 39 |
| HLA-B*08:01 | 7.24 | 10 | 18 | 12 | 29 |
| HLA-B*18:01 | 7.24 | 10 | 4 | 7 | 25 |
| HLA-C*07:02 | 11.71 | 2 | 2 | 1 | 10 |
| HLA-C*06:02 | 10.96 | 4 | 2 | 1 | 5 |
| HLA-C*07:01 | 12.03 | 0 | 0 | 0 | 0 |
| HLA-C*04:01 | 12.41 | 0 | 1 | 0 | 0 |

**Figure 5.**
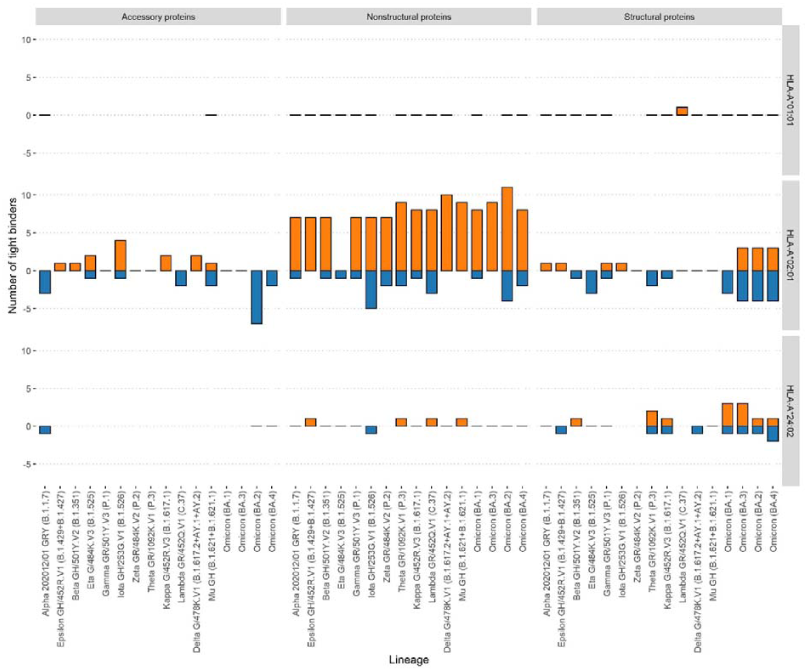
Effect of mutations in the VOC on the number of tight binders for alleles HLA-A*01:01, HLA-A*02:01, HLA-A*24:02. The orange color bars show the number of peptides that increased the affinity to 50 nM and less; the blue color bars indicate the number of peptides with a decreased affinity to more than 50 nM.

## Discussion

We conducted a comparative study of the HLA-I genotypes of symptomatic COVID-19 patients of the pandemic’s first and third waves. Genotypes of 147 patients (first wave) and 219 (third wave) were studied. We found a significant increase in the proportion of patients with obstructive pulmonary disease, obesity, and hypertension in the third wave of COVID-19. This circumstance may be associated not only with the peculiarities of the Delta variant but also with the possible population’s fatigue from complying with anti-epidemic measures, which led to the infection of risk groups that previously more strictly kept the social distancing regime (35).

We tested whether the frequency of HLA alleles can differentiate individuals from three groups: COVID-19 patients of the first wave, COVID-19 patients of the third wave, and the control group. We found that for all possible group comparisons, only one allele out of the most abundant alleles had a statistically significant odds ratio after multiple testing correction. Namely, the frequency of the HLA-A*01:01 allele in the Wave 3 group fell by half relative to the Wave 1 group. Previously, the HLA-A*01:01 allele was considered a risk allele for infection and severe course of COVID-19 in the first wave (17,36–38). At the same time, the protective role of this allele against the formation of Severe Bilateral Pneumonia caused by COVID-19 was also reported (39).

We suggested that the decrease in the frequency of HLA-A*01:01 carriers among patients of the third wave could be associated with the characteristics of the previously formed T-cell responses. It is known that up to 50% of cases of COVID-19 are asymptomatic (40,41) and lead to the formation of neutralizing antibodies and multispecific T-cells (42). While the efficiency of neutralizing antibodies decreases for VOCs (43), the formed pool of multispecific T-cells in most cases can provide an immune response regardless of viral mutations (5).

To test this hypothesis, we first analyzed the number of viral peptides interacting with the 12 most common alleles in the European population with bioinformatics methods. HLA-A*01:01 allele was one of the alleles with a moderate ability to interact with peptides of both structural and non-structural proteins of the virus. We also found that HLA-A*01:01 carriers have fewer predicted high-affinity peptides compared to the non-carriers, regardless of the wave of COVID-19.

Analysis of the confirmed immunogenic epitopes of SARS-CoV-2 according to the IEDB database showed that 10 immunoprevalent epitopes from ORF1ab and only 3 not from ORF1ab were identified for HLA-A*01:01. In turn, there were 51 immunoprevalent epitopes from ORF1ab and 59 epitopes not from ORF1ab for the HLA-A*02:01 allele. Thus, the ratio of immunoprevalent epitopes from ORF1ab for the HLA-A*01:01 allele was 3.8 times greater than for the HLA-A*02:01 allele. We additionally validated these data by analyzing the T-cell responses of 28 carriers HLA-A*01:01 and HLA-A*02:01 alleles. Concordantly with IEDB data, the true positive rates for ORF1ab epitopes were significantly higher for HLA-A*01:01 compared to the HLA-A*02:01. One of the possible explanations for these data may be the higher proportion of formed central memory CD8^+^ T-cells in HLA-A*01:01 carriers compared to the A*02:01, B*07:02, and A*24:02 carriers.

Moreover, an ORF1ab-derived HLA-A*01:01-restricted epitope TTDPSFLRGY was shown in multiple studies to induce exceptionally high frequency of T-cells (magnitude of response) in comparison to other multiple immunoprevalent epitopes (44–47). Furthermore, it was shown that the response of PBMCs of HLA-A*01:01+ convalescents to the epitopes derived from non-structural proteins was higher than to the structural proteins. This was not observed for HLA-A*01:01-convalescents (11). This further suggests high importance of ORF1ab-focused T-cell response for carriers of HLA-A*01:01.

Not a single immunoprevalent HLA-A*01:01 epitope significantly changed its presentation affinity due to mutations in the actual VOCs: Delta G/478K.V1, Omicron (BA.1 – BA. 4). At the same time, other alleles such as HLA-A*02:01 were affected by the mutations. Interestingly, HLA-A*01:01 was the only allele with a relatively high number of high-affinity peptides unaffected by the mutations. In agreement with the reports on the lower mutation rate in ORF1ab (48), high-affinity peptides from ORF1ab for all analyzed alleles were less affected by mutations compared to the rest of the proteins.

This study demonstrates for the first time the possibility of HLA-A*01:01 allele vanishment between the cohorts of symptomatic COVID-19 patients from different waves. Furthermore, the discovered characteristics of peptide presentation by HLA-A*01:01 could be important for developing vaccines to induce T-cell responses.

## Supporting information

Supplemental Tables 2 and 4. Headers Supplemental Tables 1 and 3.for

## Data Availability

All data produced in the present work are contained in the manuscript

## Acknowledgments

The research was performed within the framework of the “Creation of Experimental Laboratories in the Natural Sciences Program” at the HSE University. Funding for open access charge: Laboratory for Research on Molecular Mechanisms of Longevity, HSE University.

## Supplementary figures

**Figure S1.**
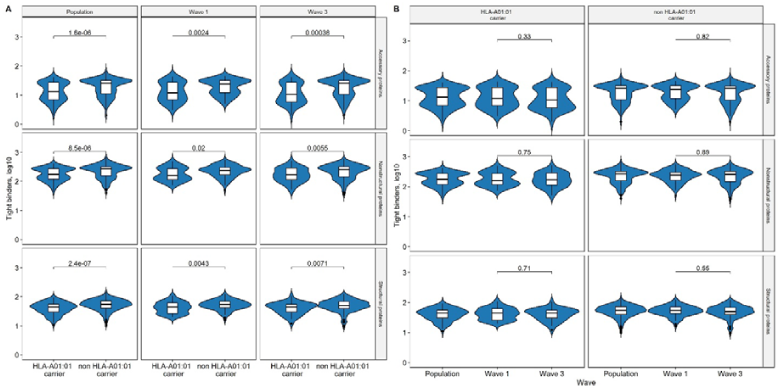
The number of unique viral peptides interacting with the sets of HLA molecules corresponding to the genotypes (affinity of no more than 50 nM). (A) Comparison of HLA-A*01:01 vs non HLA-A*01:01 carriers. (B) Comparison of Wave 1 vs Wave 3 groups.

