## Supplemental Tables 2 and 4. Headers Supplemental Tables 1 and 3.for for "HLA-A*01:01 allele vanishing in COVID-19 patients population associated with non-structural epitope abundance in CD8^+^ T-cell repertoire"

### Supplementary tables

**Table S1. HLA class I genotypes of Wave 1, Wave 3, and Control groups.**

**Table S2. The relationship of comorbidities to the risk of death from COVID-19.** OR (p-value)

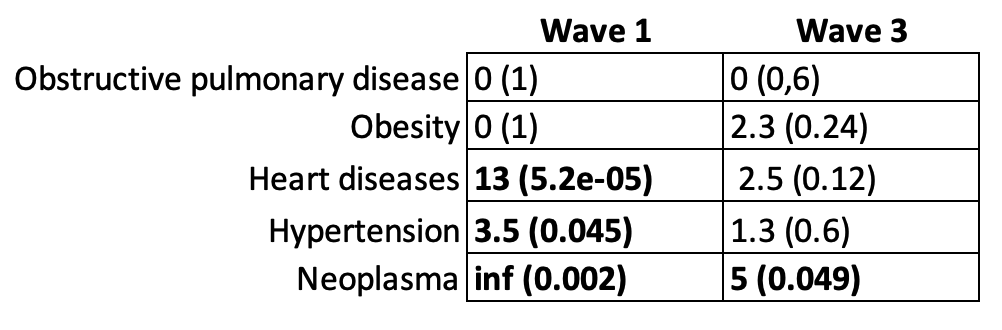

**Table S3. Association of HLA Class I Genotypes With Waves of Coronavirus Disease-19**

**Table S4. A panel of ORF1ab epitopes restricted to HLA-A*01:01 and HLA-A*02:01.**

| **Peptide** | **Location** | **Predicted binding affinity to** | |
| --- | --- | --- | --- |
|  |  | **HLA-A*01:01** | **HLA-A*02:01** |
| CTDDNALAYY | Non-structural proteins | 3 | 19896 |
| TTDPSFLGRY | Non-structural proteins | 5 | 27903 |
| DTDFVNEFY | Non-structural proteins | 6 | 33483 |
| GTDLEGNFY | Non-structural proteins | 9 | 33917 |
| PTDNYITTY | Non-structural proteins | 12 | 35372 |
| NTCDGTTFTY | Non-structural proteins | 14 | 25394 |
| HTTDPSFLGRY | Non-structural proteins | 42 | 33468 |
| YLDAYNMMI | Non-structural proteins | 221 | 2 |
| FTYASALWEI | Non-structural proteins | 5189 | 17 |
| FLLNKEMYL | Non-structural proteins | 13747 | 2 |
| YLFDESGEFKL | Non-structural proteins | 18838 | 6 |
| ALWEIQQVV | Non-structural proteins | 27002 | 5 |
| NMLRIMASL | Non-structural proteins | 29822 | 46 |
| RQLLFVVEV | Non-structural proteins | 31003 | 17 |
| KLWAQCVQL | Non-structural proteins | 33067 | 8 |
